# Newborn Screening Trial Designs: A Scoping Review Study Protocol

**DOI:** 10.1101/2025.05.22.25328131

**Authors:** Maheshi P Wijesekera, James S Bowness, Anthony J Howard, Sian Taylor-Philips, Dan Perry

## Abstract

**Background:** For policy makers to implement a screening programme, it should undergo testing to demonstrate efficacy and cost-effectiveness. When testing a screening intervention, the study design has nuances compared to conventional randomised controlled trials (RCTs). Important considerations include the timing and technique of the randomisation and consent process.

Newborn screening seeks to screen a baby for serious conditions within the first eight weeks of life. This is a broad field encompassing a variety of diseases, mainly rare conditions. There is growing interest in childhood screening among politicians in countries such as the UK, and leading organisations including the World Health Organisation (WHO). Clinical trials in newborn screening have the added complexity that consent is sought from parents to test their child. This study aims to undertake a scoping review of RCTs in newborn screening to summarise the trial designs, with particular reference to randomisation and consent processes.

**Methods:** The search will be carried out on OVIDMEDLINE, EMBASE, CINAHL and CENTRAL databases. Clinical trials registries, approved by the International Committee of Medical Journal Editors (ICMJE) and the WHO clinical trial registry, will be searched for any studies registered. Additionally in those studies that are included any citing or cited literature will be searched for any relevant studies.

**Discussion:** This study is the first scoping review of its kind. Synthesis of the available evidence will enable investigators to better consider the nuances of undertaking screening trials among newborns when designing future evaluations of screening programmes in children.

## Introduction

Newborn screening is a cornerstone of public health aimed at the early identification of rare diseases. It aims to screen infants in their first eight weeks of life and allows timely intervention to reduce morbidity and mortality.^1^ By detecting conditions before symptoms develop, screening programs provide opportunities for early treatment, improving long-term health outcomes and enabling families to make informed decisions. However, despite the potential benefits, the design and implementation of newborn screening research trials present unique methodological and ethical challenges.^2^

Screening differs from therapeutic interventions, as it is a secondary prevention strategy designed to identify disease in asymptomatic populations. For a screening programme to be effective, it must meet criteria such as those established by Wilson and Jungner, ensuring that early intervention leads to meaningful health improvements and that the benefits outweigh potential harms.^3^ Many countries have integrated newborn screening into national programs, but the complexity of screening in newborns especially for rare, inherited diseases raises distinct challenges in trial design, randomisation methods, and consent processes.^4,5^

### Challenges in Conducting Screening Trials

Before implementation into routine practice, a screening test should be rigorously evaluated in a trial to assess its efficacy and impact on health outcomes.^4,6^ Randomised controlled trials (RCTs) are considered the gold standard of evidence based medicine; however, practical and ethical constraints often limit their feasibility in screening research. Unlike therapeutic trials, screening trials require large cohorts of healthy participants, long term follow up, and high retention rates, making them particularly challenging.^6^

The key ethical barrier in newborn screening trials lies in establishing a control group that does not undergo screening when the efficacy of the screening has yet to be established. While robust evidence is essential to justify the population-wide implementation of a screening program, purposefully withholding screening when early diagnosis and treatment may alter the natural history of the disease raises concerns related to non-maleficence and the potential failure to uphold a newborn’s best interests.^7,8^ This highlights the ethical dilemma of achieving scientific validity against the obligation of protecting vulnerable populations from avoidable harm. The use of a control arm employing the next-best screening method (such as an alternative screening test or using selective screening versus universal screening) does not necessarily resolve this concern, as some infants may still miss out on the opportunity of early treatment if one method proves inferior. Another significant ethical issue relates to informed consent. While informed consent is a requirement for participation in research, it is debatable whether parents truly understand the implications particularly the severity of rare conditions, the potential benefits of early detection, or the limitations such as not receiving any immediate benefit for their child.^2^ This effect amplifies when consent is sought at population-level, potentially diluting the quality and consistency of information provided by recruiters.^9^ For example variability in communicating risks, test implications, and follow-up procedures.

Therefore traditional trial models may not be fully appropriate in this context, as the process of obtaining informed consent can both hinder recruitment and inadvertently exclude infants from receiving potentially beneficial interventions. Adopting alternative trial designs and innovative consent frameworks may therefore be necessary to ethically and effectively conduct newborn screening research.^4,10,11^

### Key Considerations in Screening Trial Methodology

Three major methodological considerations influence the design of screening trials:

1. **Trial designs** – Traditional screening trials randomise participants into screened and non-screened groups, but alternative designs have emerged to improve recruitment and feasibility. Patient-preference designs, quasi experimental study designs and observational studies have been employed when RCTs are impractical.^4,12,13^
2. **Randomisation methods**-Given the difficulty of randomising healthy individuals, innovative approaches such as the Zelen design, Trial Within Cohort (TWiCs), comprehensive cohort design, and Wennberg’s design have been used to balance scientific rigour with participant acceptability.^14–16^
3. **Consent processes**-Informed consent can be pre-randomisation (before allocation) or post-randomisation (after allocation), each with distinct ethical and practical implications. In newborn screening, consent challenges are amplified by the need for parental decision-making within a short postnatal window.^2^

### Newborn Screening as a Complex Intervention

The UK Medical Research Council (MRC) defines complex interventions as those with multiple interacting components that influence real-world decision-making.^17^ Newborn screening fits this definition, given its reliance on multifaceted policies, diverse screening modalities, and ethical considerations surrounding consent, autonomy, and privacy. Furthermore, screening trials do not merely test an intervention; they interact with broader public health policies, healthcare systems, and societal expectations, making their evaluation inherently complex.^18^

In the UK, newborn screening includes the Newborn and Infant Physical Examination (NIPE), newborn hearing screening, and the newborn blood spot test, which screens for conditions such as cystic fibrosis, sickle cell disease, and metabolic disorders (see Table 1).^19^ The UK National Screening Committee (UK NSC) requires high-quality evidence, preferably from RCTs, to support the introduction of new screening tests.^20^ However, delivering traditional RCTs in newborn screening is often impractical due to the rarity of conditions, making high-quality observational and alternative trial designs essential.^8,21^

**Table 1:** UK Newborn screening tests.

| Newborn and Infant Physical Examination (NIPE) | Blood spot test |
| --- | --- |
| Congenital cardiac disease | Sickle cell disease |
| Congenital cataracts | Cystic fibrosis |
| Developmental dysplasia of the hip | Congenital hypothyroidism |
|  | Inherited metabolic conditions:<br>- Phenylketonuria (PKU)<br>- Medium-chain acyl-CoA dehydrogenase deficiency (MCADD)<br>- Maple syrup urine disease (MSUD)<br>- Isovaleric acidaemia (IVA)<br>- Glutaric aciduria type 1 (GA1)<br>- Homocystinuria (pyridoxine unresponsive) (HCU)<br>- Tyrosinaemia |

### A screening trial demonstrating the challenges

The Wisconsin cystic fibrosis neonatal screening project was a pivotal example of a large scale newborn screening trial, highlighting key methodological and ethical challenges.^22^ Initiated in 1985, this RCT screened over 650,000 newborns to assess whether early detection of cystic fibrosis improved population health outcomes. Participants were randomly assigned to either a screened group, receiving cystic fibrosis testing, or a control group, where screening results were withheld until symptoms developed. While the trial provided compelling evidence that early diagnosis led to long-term health benefits, it faced significant ethical scrutiny; particularly regarding the control group’s decision to withhold the test results from the control group. The legal and ethical challenges arising from this study underscores the importance of robust consent processes, transparent communication with parents, and careful consideration of the treatment of the control group in newborn screening trials. The lessons from the Wisconsin trial remain highly relevant in shaping future screening trial designs and ensuring ethical integrity in neonatal research.^2,23^

### The Need for a Scoping Review

Despite the widespread implementation of newborn screening, there is no comprehensive synthesis of the trial designs, randomisation strategies, and consent processes used in screening trials. A preliminary search (conducted on 14th January 2025) of databases including the Joanna Briggs Institute, Cochrane Library, and PROSPERO found no existing scoping or systematic reviews registered on this topic. While systematic reviews have evaluated specific conditions, they have not examined the specific methodological considerations or ethical challenges in newborn screening research.^24–29^

Given the broad scope of newborn screening and the diverse methodologies used in evaluating these programs, a scoping review is the most appropriate approach. Scoping reviews differ from systematic reviews by mapping key concepts, study designs, and methodological challenges in a research area rather than focusing on a narrow clinical question.^30^ This review aims to:

1. Identify the range of trial designs used in newborn screening research which employ a randomised study design
2. Explore the randomisation methods employed
3. Evaluate consent processes, and ethical models employed and their implications
4. Extract key lessons to inform future newborn screening trial design

Currently there is a need to undertake a good quality evidence to demonstrate if newborn screening is required for conditions such as developmental dysplasia of the hip and critical congenital cardiac disease in the UK NIPE. Therefore synthesising available evidence, this review will contribute to the future development of ethically sound, scientifically rigorous, and practically feasible screening trials, ultimately enhancing the effectiveness of newborn screening programs worldwide.

## Methods

### Protocol and Registration

This protocol was developed using the conduct guidance from the Joanna Briggs Institute (JBI) and will be reported based on the Preferred Reporting Items for Systematic Reviews and Meta Analyses Extension for Scoping Reviews (PRISMA ScR).^31–33^ The protocol will be registered with medRxiv (https://www.medrxiv.org) prior to data extraction, as shown in Appendix 1 and any deviations from the protocol will be clearly explained in the subsequent publication.

### Scoping review question

What trial designs, randomisation methods and consent processes have been undertaken for newborn screening, and what can we learn from this to inform future trials?

### Inclusion criteria

#### Population

All studies evaluating the clinical and/or cost effectiveness of newborn screening performed amongst infants under the age of 8 weeks old. Only data from human studies will be included. Studies should primarily screen infants <8 weeks old though we recognise that in some instances older children may also be screened outside the primary target population.

#### Concept

The data assessed will relate to the study designs with respect to the trial design, randomisation techniques and consent processes. Consent process will be assessed based if consent was active or passive and based on timing of randomisation (i.e. pre randomisation or post randomisation), individual or consent waiver used in screening studies. Only studies investigating a screening intervention in the newborn population will be included. Any study analysing a population level screening programme will also be included.

#### Context

Studies evaluating screening programmes or screening tests.

#### Types of sources

All published research, including academic publications and protocols, will be considered. This scoping review will consider experimental study designs that are prospective in nature. This includes RCTs, with cluster or pragmatic study designs. Additionally, TWiCs and patient preference trials will be considered if they include a randomisation element within the study design.

There will be no year limit to the searches. The search will be limited to only those published in the English language.

#### Exclusion criteria

Any non-interventional and quasi experimental study designs such as pre-post intervention designs with a non-equivalent control group or interrupted time series study designs will be excluded. If an RCT was performed as a decision aid to improve up take of screening this would be excluded.

#### Search Strategy

The lead investigator developed the search strategy and reviewed/ modified by a medical librarian.

A three-step search strategy will be incorporated. Given the iterative nature of scoping reviews, the search will be carried out initially on one databases (EMBASE) using search terms demonstrated in table 2. Searches will be limited to subjects scanned to human only, and limited to language (English only studies). The search will exclude any case reports or review articles. The title and abstracts will be screened for articles to be included. The index words of included manuscripts will be reviewed and added to the search strategy if already not included. Any additional data for extraction will be supplemented to the data extraction table, appendix 1.The full search will then be executed. The search will be conducted out on Medline, CINAHL (via the NICE Healthcare Databases Advanced Search interface), and Cochrane Central Register of Controlled Trials (CENTRAL).

**Table 2:** Search terms.

| Concept |  | MeSH terms |
| --- | --- | --- |
| #1 | “Newborn” “neonat*” OR “infant” | newborn |
| #2 | “screening” | Mass screening |
|  |  | neonatal screening |
| #3 | “Clinical trial” OR “random* trial” OR “controlled trial” OR “RCT” OR “cmRCT” OR “Cohort Multiple Randomised Controlled Trial” OR “TWICS” OR “trial within cohort” OR Zelen | Clinical trial<br>Randomised controlled trial |
| #4<br>*Exclusion terms* | “Antenatal” OR “pregnancy” OR “gestational” OR “fetal” OR “foetal” OR “prenatal” OR “perinatal” OR “placenta” OR “embryo” |  |

*The search combination would include #1 AND #2 AND #3 AND NOT #4. AND NOT #5*

The registry search will include databases approved by International Committee of Medical Journal Editors (ICMJE). (www.anzctr.org.au ; www.clinicaltrials.gov ; www.ISRCTN.org ; www.umin.ac.jp.ctr ; www.trialregister.nl ; https://eudract.ema.europa.eu via the EU Clinical Trials Register (www.clinicaltrialsregister.eu). In addition, the World Health Organisation (WHO) clinical trials registry (http://www.who.int/ictrp/search/en) will be searched for any studies registered.

Cited literature in, and literature citing, the included studies will be scrutinised for relevance. Any relevant sources will be included for subsequent data extraction.

Grey literature will not be included given the focus of this study is clinical trials and it is expected that the results or protocols will be published.

### Data Extraction

#### Title review

Once results from all sources have been collated, two investigators will screen titles. The initial screen will be to screen inclusion in the study, with titles used to determine if they are related to newborn screening. Any identified duplications will be removed. In cases of discrepancy, a third investigator will review the title to adjudicate. If the title is ambiguous the source will be included in the abstract review.

#### Abstract Review

An investigator will review the abstract of all included sources to consider for a full-text review. Sources will be included if the abstract/summary relates to screening in newborns undertaking an experimental study.

#### Extraction

Full texts will be reviewed to determine the suitability for data extraction. The study designs will be scrutinised to extract data on trial design, randomisation and consent processes used. The extracted data will be electronically charted using the Table in Appendix 1. Data extraction will be considered an iterative process, such that if any further variable becomes apparent, this too will be added to the table for data extraction.^34^

The search and screening process will be shown in a flowchart that details the number of publications considered at each stage. Data extraction will initially be conducted by a single author, with another checking over the data extraction to ensure full scrutiny of each source. To ensure uniformity and minimise any error associated with data extraction, piloting of data extraction will be undertaken among the review team.^34^ If any discrepancies occur these will be resolved through discussion with a third reviewer (JB).

If a study protocol or publication has insufficient information, the corresponding author will be contacted to obtain any additional information.

#### Data presentation

Extracted data will be summarised using tables and presented in a narrative format. Should there be sufficient data on the topic, a graphical method incorporating plots will be utilised to present data.

## Discussion

The proposed scoping review aims to summarise the available evidence on the study designs in newborn screening and will expand on the techniques used by each source using a narrative approach.

Where sufficient data is identified, the data is likely to inform the optimal trial design for newborn screening trials. It is hoped that this review can serve as a summary for future applications within paediatric research.

## Limitations

Despite the systematic search strategy, we anticipate some challenges. We recognise the possibility of publication bias, where trials have been initiated but unable to be completed due to challenges in recruitment which may fail to identify the challenges faced by types of study. However, these trials should be identified through published protocols, and authors would be contacted for any additional information not presented. To keep the search manageable, the review will be limited to studies published in English. Limiting the search to English-language studies was justified based on the predominance of high-quality clinical research published in English-language journals, nevertheless, we acknowledge the potential shortcomings of this approach.

Scoping reviews follow an iterative process and are exploratory in nature. Consequently, the complete breadth of relevant literature may not be apparent at the outset, which may need the search strategy to be modified as the study progresses.^34^ We anticipate that some studies will lack clarity, failing to clearly include descriptions of the patient populations or and research methods used. This is a recognised limitation and we will seek to contact the corresponding authors to seek further information where required.^30^

Our definition of newborn screening expands to 8 weeks old, which is consistent with the UK newborn screening time frame. Terminology in other countries, such as the United States, can be different (e.g., newborn screening here takes place in the first two weeks of life). This may affect the generalisability of our findings.

## Data Availability

Given that this is a study protocol no data is available in the manuscript

## Appendices

### Appendix 1

#### Data extraction sheet

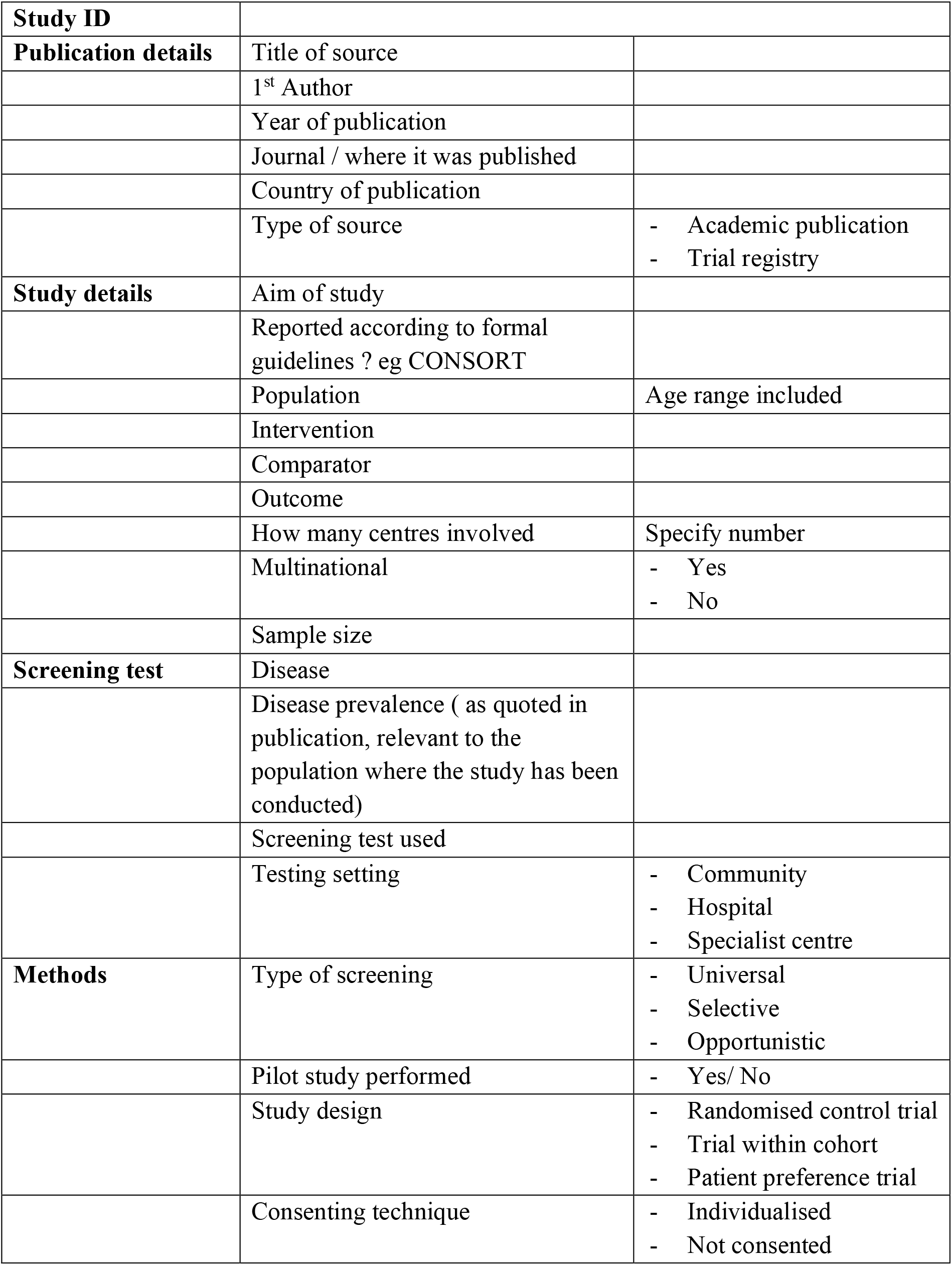
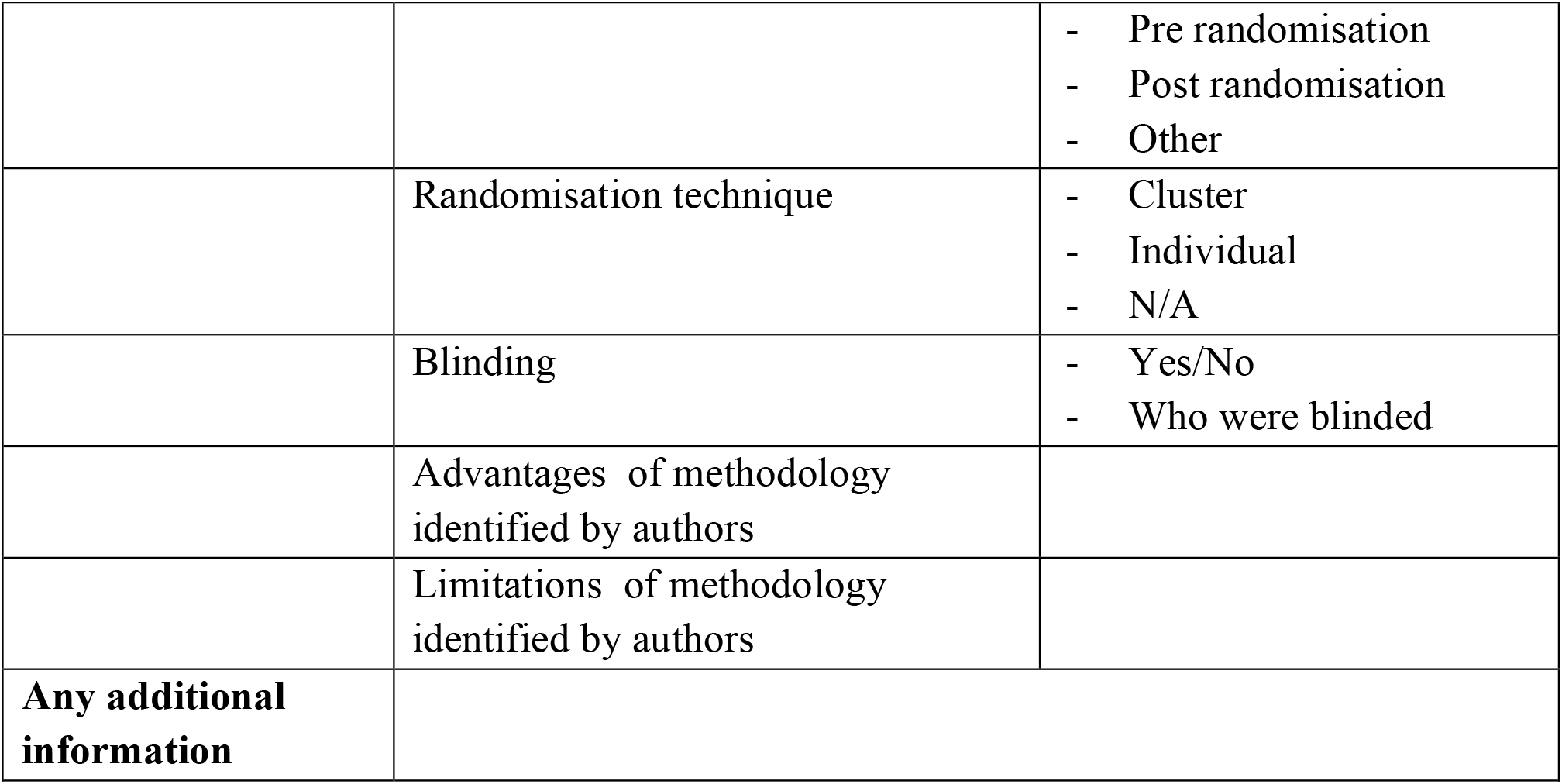

## Notes

### Competing Interest Statement

JSB works for GE Healthcare, and previously consulted/worked for AutonomUS Medical Technologies and Intelligent Ultrasound. He has received speaker fees from the Belgian Association of Regional Anaesthesia, as well as King Fiasal Specialist Hospital and Research Centre
STP: Member of the UK National Screening Committee, NIHR research professor
DP: national clinical advisor for hip screening to the NHS England antenatal and newborn programme advisory group, NIHR research professor

### Funding Statement

This study did not receive any funding

